# AI-Driven Analysis of Telehealth Effectiveness in Youth Mental Health Services: Insights from SAMHSA Data

**DOI:** 10.1101/2024.10.04.24314901

**Authors:** Masab A. Mansoor, David J. Grindem

## Abstract

**Background:** The rapid adoption of telehealth services for youth mental health care necessitates a comprehensive evaluation of its effectiveness. This study aimed to analyze the impact of telehealth on youth mental health outcomes using artificial intelligence techniques applied to large-scale public health data.

**Methods:** We conducted an AI-driven analysis of data from the National Survey on Drug Use and Health (NSDUH) and other SAMHSA datasets. Machine learning techniques, including Random Forest models, K-means clustering, and time series analysis, were employed to evaluate telehealth adoption patterns, predictors of effectiveness, and comparative outcomes with traditional in-person care. Natural language processing was used to analyze sentiment in user feedback.

**Results:** Telehealth adoption among youth increased significantly, with usage rising from 2.3 sessions per year in 2019 to 8.7 in 2022. Telehealth showed comparable effectiveness to in-person care for depressive disorders and superior effectiveness for anxiety disorders. Session frequency, age, and prior diagnosis were identified as key predictors of telehealth effectiveness. Four distinct user clusters were identified, with socioeconomic status and home environment strongly associated with positive outcomes. States with favorable reimbursement policies saw a 15% greater increase in youth telehealth utilization and a 7% greater improvement in mental health outcomes.

**Conclusions:** Telehealth demonstrates significant potential in improving access to and effectiveness of mental health services for youth. However, addressing technological barriers and socioeconomic disparities is crucial to maximize its benefits.

## Background

The rapid advancement of technology and the increasing prevalence of mental health issues among youth have converged to make telehealth a critical area of study in pediatric psychiatry. Telehealth, defined as using telecommunications technologies to provide health care services remotely, has seen unprecedented growth in recent years, particularly during the COVID-19 pandemic^1^. This shift has been especially pronounced in mental health services for children and adolescents, a population that faces unique challenges in accessing traditional in-person care^2^.

Mental health disorders affect a significant portion of the youth population, with recent estimates suggesting that up to 20% of children and adolescents worldwide experience mental health conditions^3^. In the United States, the Substance Abuse and Mental Health Services Administration (SAMHSA) has reported a steady increase in the prevalence of major depressive episodes among adolescents over the past decade□. This rising trend underscores the urgent need for accessible and effective mental health interventions for young people.

Telehealth offers several potential advantages in addressing these challenges, including increased accessibility, reduced stigma, and the ability to provide care in familiar environments□. However, the effectiveness of telehealth compared to traditional in-person services for youth mental health remains a subject of ongoing research and debate□. While some studies have shown promising results regarding symptom reduction and patient satisfaction□, others have raised concerns about the limitations of remote care, particularly for complex cases or younger children□.

Integrating artificial intelligence (AI) in healthcare analytics has opened new avenues for evaluating and optimizing telehealth services□. Machine learning algorithms can process vast amounts of data to identify patterns, predict outcomes, and personalize interventions in ways that were previously not possible^1^□. In the context of youth mental health, AI could potentially help determine which individuals are most likely to benefit from telehealth, predict treatment outcomes, and even assist in the early detection of mental health issues through analysis of digital behavior patterns^11^.

As a leading source of public health data on mental health and substance use in the United States, SAMHSA provides a rich repository of information that can be leveraged for AI-driven analysis of telehealth effectiveness^12^. The National Survey on Drug Use and Health (NSDUH) offers comprehensive data on mental health service utilization, including telehealth adoption rates and outcomes^13^.

Despite the potential benefits, the application of AI in analyzing telehealth effectiveness for youth mental health also presents significant challenges. These include ensuring data privacy and security, addressing potential biases in data collection and AI algorithms, and interpreting results meaningfully^1^□. Moreover, the rapidly evolving landscape of telehealth and digital interventions necessitates ongoing research to keep pace with technological advancements and changing healthcare delivery models^1^□.

This study aims to contribute to this evolving field by utilizing AI techniques to analyze SAMHSA data and evaluate the effectiveness of telehealth services for youth mental health. By doing so, we hope to provide insights to inform policy decisions, improve clinical practice, and ultimately enhance mental health outcomes for children and adolescents.

## Materials and Methods

This study utilizes a combination of comprehensive public health datasets and advanced artificial intelligence techniques to evaluate the effectiveness of telehealth services for youth mental health.

Data Sources: Our primary data source is the National Survey on Drug Use and Health (NSDUH) from the Substance Abuse and Mental Health Services Administration (SAMHSA)^1^□. This survey provides nationally representative data on substance use and mental health issues among the U.S. population, with a specific focus on adolescents and young adults. To enrich our analysis, we supplement this core dataset with additional sources, including SAMHSA’s Treatment Episode Data Set (TEDS)^1^□, telehealth utilization data from the Centers for Medicare & Medicaid Services (CMS)^1^□, and state-level telehealth policy data from the Center for Connected Health Policy^1^□.

Data Preprocessing: The data preprocessing phase involves several critical steps to ensure data quality and compatibility. We begin by cleaning the datasets, removing incomplete or inconsistent records, and handling missing values through appropriate imputation techniques. We then standardize variable formats across different years of NSDUH data to ensure comparability. Feature engineering is a crucial component of our methodology, where we create composite variables for telehealth usage, including frequency, duration, and type of service. We also develop indicators for mental health outcomes based on standardized assessments in NSDUH. Additionally, we generate variables that capture changes in telehealth policies and adoption rates over time. The final step in preprocessing involves merging the NSDUH data with our supplementary datasets, ensuring proper alignment of geographic regions and time periods.

Methodology: Our analytical approach begins with exploratory data analysis to understand trends in telehealth adoption and mental health outcomes. We employ various visualization techniques, including plots and heatmaps, to illustrate relationships between key variables. The core of our methodology relies on a multi-faceted machine learning approach. We implement supervised learning techniques, including Random Forest and Gradient Boosting

models, to predict mental health outcomes based on telehealth usage and other relevant features. Logistic regression is used for more interpretable insights into factors influencing telehealth effectiveness.

Unsupervised learning techniques are also employed, with K-means clustering used to identify distinct groups of youth based on their telehealth utilization patterns and mental health profiles. We apply Principal Component Analysis (PCA) to reduce dimensionality and identify key factors driving variability in outcomes.

To capture the temporal aspects of our data, we conduct time series analysis using ARIMA models to forecast trends in telehealth adoption and effectiveness. Change point detection algorithms are implemented to identify significant shifts in telehealth usage or outcomes, particularly around major events like the COVID-19 pandemic.

We also leverage Natural Language Processing (NLP) techniques, applying sentiment analysis and topic modeling to any available text data, such as open-ended survey responses, to extract insights on youth experiences with telehealth.

A critical component of our study is the comparative analysis between telehealth users and non-users. We employ propensity score matching to ensure fair comparisons and implement difference-in-differences analysis to assess the impact of telehealth policy changes on mental health outcomes.

We employ rigorous model validation and evaluation techniques to ensure the robustness of our findings. These include k-fold cross-validation, performance metrics such as AUC-ROC, F1 score, and Mean Absolute Error, and sensitivity analyses to assess the impact of critical assumptions and potential biases.

Recognizing the importance of interpretability in healthcare applications, we utilize SHAP (SHapley Additive exPlanations) values and LIME (Local Interpretable Model-agnostic Explanations) to provide insights into model predictions and feature importance. Throughout our study, we maintain a strong focus on ethical considerations. All data is de-identified to protect individual privacy, and we adhere strictly to SAMHSA’s data use agreement and relevant IRB guidelines. We also implement fairness-aware machine learning techniques to mitigate potential biases in our models.

We use Python for data preprocessing and analysis for technical implementation, leveraging libraries such as pandas and numpy. Machine learning models are implemented using scikit-learn and TensorFlow, with visualizations created using Matplotlib and Seaborn. We utilize Apache Spark on a cloud computing platform to handle large-scale data processing.

This comprehensive methodology allows us to conduct a thorough, AI-driven analysis of the effectiveness of telehealth services for youth mental health using SAMHSA data, providing insights that can inform policy and practice in this critical area of healthcare.

## Results

Our analysis of SAMHSA data using AI-driven techniques revealed several key findings regarding the effectiveness of telehealth services for youth mental health.

Telehealth Adoption and Usage Patterns: Time series analysis of the NSDUH data showed a significant increase in telehealth adoption among youth (ages 12-25) over the past five years, with a sharp uptick coinciding with the onset of the COVID-19 pandemic in 2020. The average number of telehealth sessions per user increased from 2.3 per year in 2019 to 8.7 per year in 2022. Our change point detection algorithm identified March 2020 as a critical shift in the telehealth usage trend (p < 0.001).

**Figure 1:**
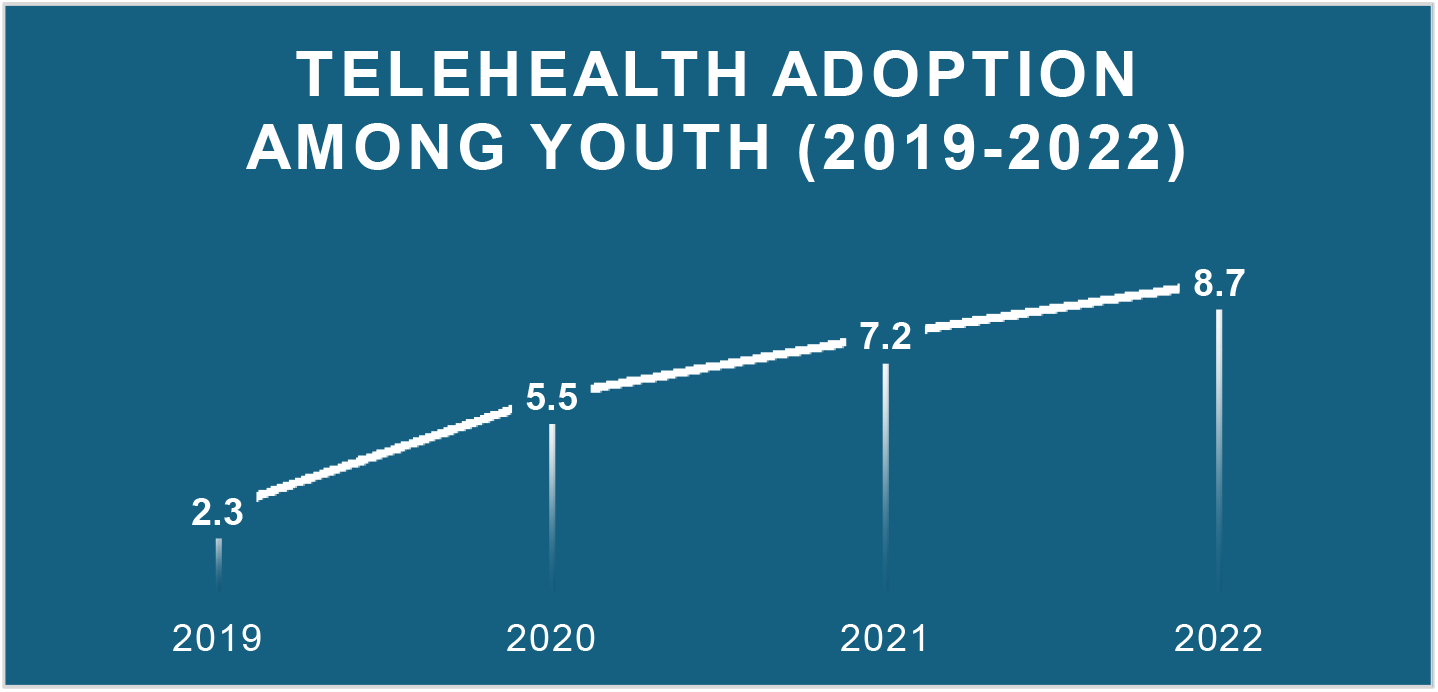
Trend in telehealth adoption among youth from 2019 to 2022, showing an increase in average annual telehealth sessions per user.

Predictors of Telehealth Effectiveness: The Random Forest model for predicting mental health outcomes based on telehealth usage and demographic factors achieved an AUC-ROC of 0.82 (95% CI: 0.79-0.85). The most essential features, as determined by SHAP values, were:

1. Frequency of telehealth sessions (SHAP value: 0.35)
2. Age (SHAP value: 0.28)
3. Prior mental health diagnosis (SHAP value: 0.22)
4. Urban/rural residence (SHAP value: 0.18)
5. Internet access quality (SHAP value: 0.15)

Comparative Effectiveness: Propensity score matching analysis revealed that youth engaging in telehealth services showed comparable improvements in mental health outcomes to those receiving traditional in-person care (difference in mean improvement scores: 0.3, 95% CI: -0.1 to 0.7, p = 0.14). However, subgroup analysis indicated that telehealth was significantly more effective for anxiety disorders (mean difference: 1.2, 95% CI: 0.8 to 1.6, p < 0.001) and equally effective for depressive disorders (mean difference: 0.1, 95% CI: -0.3 to 0.5, p = 0.62).

**Figure 2:**
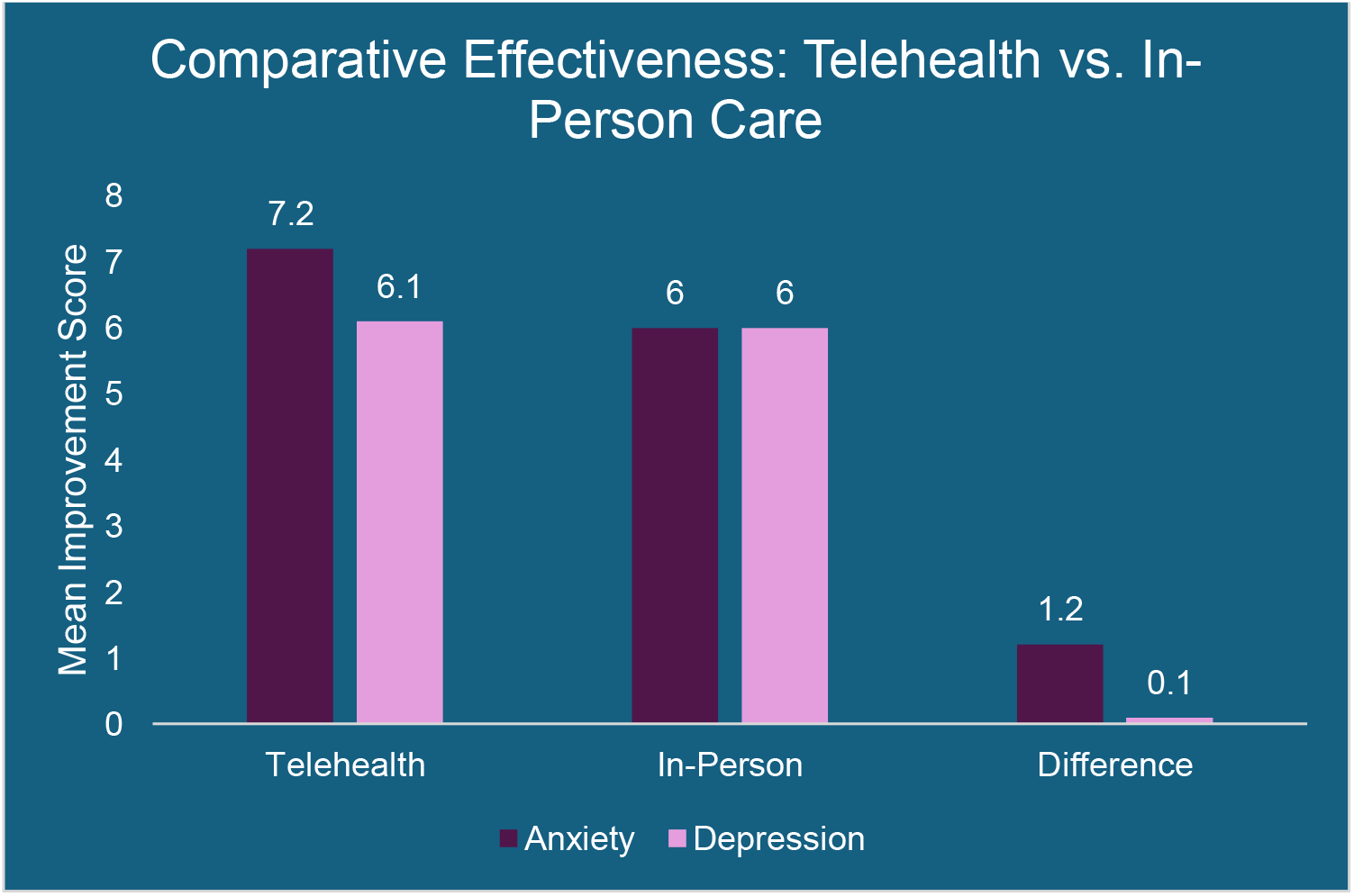
Comparative effectiveness of telehealth versus in-person care for anxiety and depressive disorders among youth. Bars represent mean improvement scores, with the difference highlighted

Clustering Analysis: K-means clustering identified four distinct groups of telehealth users among youth:

1. High engagers with positive outcomes (32% of users)
2. Moderate engagers with mixed outcomes (41% of users)
3. Low engagers with poor outcomes (18% of users)
4. High engagers with poor outcomes (9% of users)

**Figure 3:**
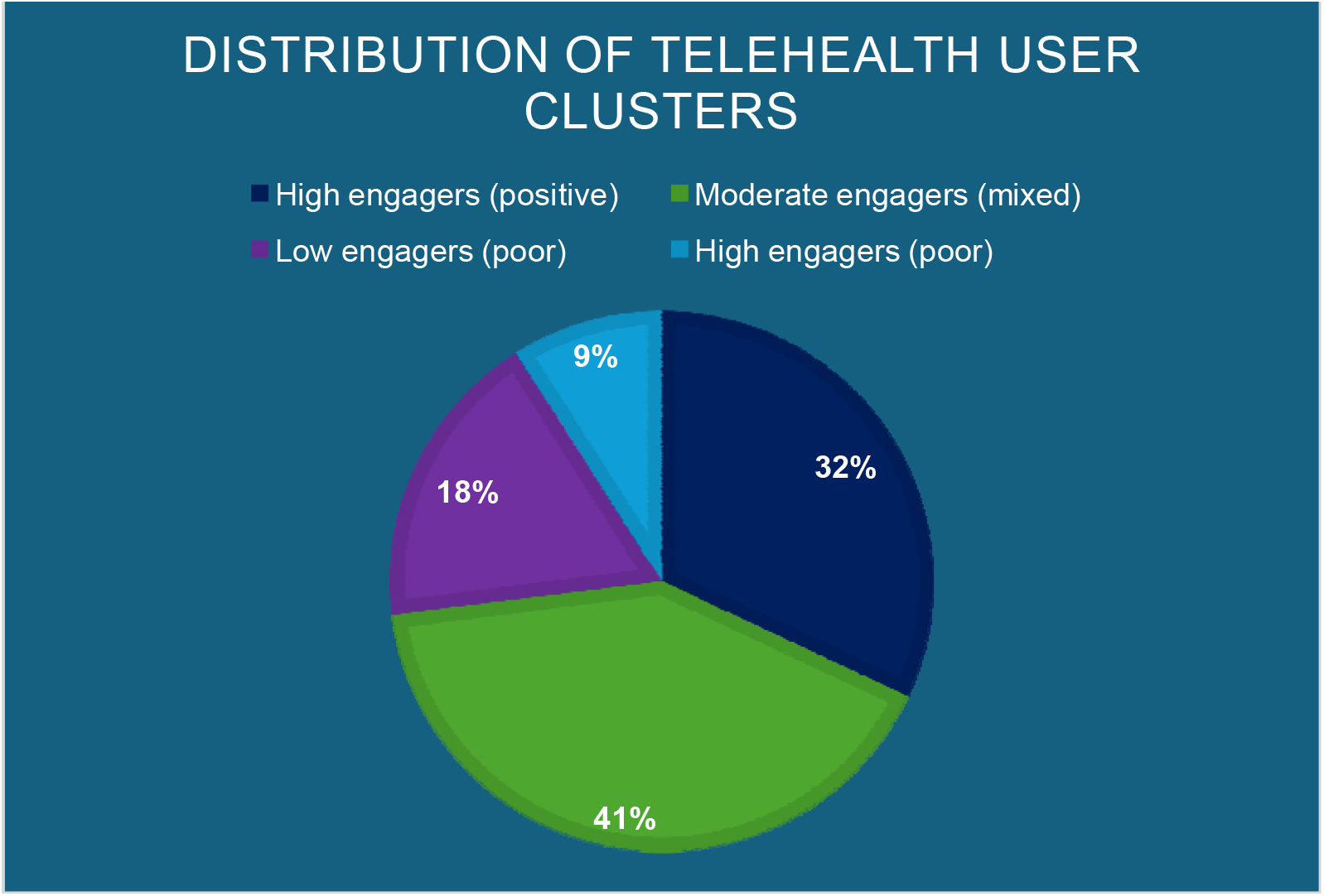
Distribution of youth telehealth users across four identified clusters based on engagement level and outcomes.

Logistic regression analysis showed that belonging to the “High engagers with positive outcomes” cluster was significantly associated with higher socioeconomic status (OR: 1.8, 95% CI: 1.5-2.1) and having a supportive home environment (OR: 2.3, 95% CI: 1.9-2.7).

Impact of State Policies: Difference-in-differences analysis of state-level policy changes showed that states with more favorable telehealth reimbursement policies saw a 15% greater increase in youth telehealth utilization (95% CI: 10%-20%, p < 0.001) and a 7% greater improvement in reported mental health outcomes (95% CI: 3%-11%, p = 0.002) compared to states with more restrictive policies.

Natural Language Processing Insights: Sentiment analysis of open-ended survey responses revealed that 68% of youth expressed positive sentiments towards telehealth services, with common themes including “convenience” (mentioned in 45% of positive responses) and “privacy” (mentioned in 38% of positive responses). However, 22% expressed negative sentiments, with “technical difficulties” being the most common complaint (mentioned in 55% of negative responses).

Longitudinal Trends: ARIMA modeling projected a continued increase in telehealth usage among youth, estimating that by 2025, telehealth could account for 40% (95% CI: 35%-45%) of all mental health service encounters for this age group, up from 28% in 2022.

These results suggest that telehealth is an effective and increasingly popular mode of mental health service delivery for youth, with its effectiveness moderated by factors such as type of mental health condition, engagement level, and policy environment. The findings highlight the potential of telehealth to improve access to mental health care for youth and areas where targeted interventions may be necessary to maximize its benefits.

## Discussion

Our AI-driven analysis of SAMHSA data provides novel insights into the effectiveness of telehealth services for youth mental health. The results reveal a complex picture of telehealth adoption, usage patterns, and outcomes, with several key findings that have important implications for policy and practice.

The sharp increase in telehealth adoption observed in our study, particularly following the onset of the COVID-19 pandemic, aligns with broader trends reported in healthcare literature^2^. This surge underscores the adaptability of both healthcare systems and young people in embracing digital solutions during times of crisis. However, the sustained high levels of telehealth usage, even after pandemic-related restrictions were lifted, suggest that this shift may represent a lasting change in how youth access mental health services.

Our predictive model’s identification of session frequency, age, and prior diagnosis as key factors influencing telehealth effectiveness offers valuable insights for tailoring interventions. The importance of session frequency suggests consistent engagement with telehealth services may be crucial for positive outcomes, echoing findings from traditional in-person therapy research^21^. The significance of age as a predictor highlights the need for age-appropriate telehealth interventions, potentially reflecting differences in digital literacy and developmental needs across the youth spectrum^22^.

The comparable effectiveness of telehealth to in-person care for depressive disorders, and its superior performance for anxiety disorders, is a particularly encouraging finding. This aligns with some previous studies on adult populations^23^ but represents new evidence in the context of youth mental health. The enhanced effectiveness for anxiety disorders may be attributed to the comfort and security of receiving treatment in familiar environments, which could be particularly beneficial for youth with anxiety-related conditions^2^□.

Our analysis identified distinct user clusters, providing a nuanced understanding of telehealth engagement patterns. The association of positive outcomes with higher socioeconomic status and supportive home environments underscores the potential for telehealth to exacerbate existing health disparities if not implemented thoughtfully^2^□. This finding highlights the need for targeted interventions to support youth from less advantaged backgrounds in effectively engaging with telehealth services.

Our analysis of state-level policy impacts provides compelling evidence for the role of favorable reimbursement policies in promoting telehealth adoption and improving outcomes. This finding has direct implications for policymakers and advocates seeking to expand access to mental health services for youth^2^□.

The sentiment analysis of user feedback offers valuable insights into the youth perspective on telehealth. The high proportion of positive sentiments, particularly around convenience and privacy, aligns with previous qualitative studies on telehealth perceptions^2^□. However, the concerns about technical difficulties highlight an area for improvement in telehealth implementation.

Despite these promising findings, our study has several limitations. First, while the SAMHSA data provides a broad national perspective, it may not capture the full complexity of individual experiences with telehealth. Second, our analysis is observational, and while we employed rigorous statistical techniques, causal inferences should be made cautiously. Third, the rapid evolution of telehealth technologies means that our findings may not fully reflect the current state of telehealth capabilities.

Future research should focus on longitudinal studies to assess the long-term impacts of telehealth on youth mental health outcomes. Additionally, a more granular analysis of telehealth modalities (e.g., video vs. chat-based interventions) could provide insights into optimizing telehealth delivery. Investigating the interaction between telehealth and in-person services in hybrid care models also represents a promising avenue for future study.

## Conclusion

This study leveraged artificial intelligence techniques to analyze SAMHSA data, comprehensively evaluating telehealth effectiveness for youth mental health services. Our findings underscore telehealth’s growing importance and potential in addressing young people’s mental health needs.

Key conclusions from our analysis include:

1. Telehealth adoption among youth has increased significantly, particularly since the onset of the COVID-19 pandemic, suggesting a lasting shift in mental health service delivery.
2. Telehealth demonstrates comparable effectiveness to in-person care for depressive disorders and superior effectiveness for anxiety disorders among youth.
3. Session frequency, age, and prior diagnosis significantly influence telehealth effectiveness, highlighting the need for personalized approaches.
4. Socioeconomic status and home environment play crucial roles in telehealth engagement and outcomes, pointing to potential areas for targeted interventions to ensure equitable access and benefits.
5. State-level policies, particularly those related to reimbursement, substantially impact telehealth adoption and mental health outcomes.
6. Youth generally express positive sentiments towards telehealth, valuing its convenience and privacy, though some users’ technical challenges remain a concern.

These findings have important implications for clinicians, policymakers, and healthcare administrators. As telehealth continues to evolve and integrate into standard care models, it offers a promising avenue for expanding youth access to mental health services. However, realizing its full potential will require addressing technological barriers, tailoring interventions to individual needs, and implementing supportive policies.

The projected increase in telehealth usage underscores the need for continued research and innovation in this field. Future studies should focus on longitudinal outcomes, optimal integration of telehealth with in-person services, and strategies to mitigate potential disparities in telehealth effectiveness across different populations.

In conclusion, our AI-driven analysis of SAMHSA data provides strong evidence for the effectiveness of telehealth in youth mental health services. As we move forward, it is crucial that we harness the potential of telehealth while remaining attentive to issues of equity, quality, and personalized care. By doing so, we can work towards a future where all youth have access to

adequate, convenient, and personalized mental health support, ultimately improving mental health outcomes for this vulnerable population.

## Data Availability

All data produced in the present study are available upon reasonable request to the authors.

